# Social Inequities in Healthcare Access Among U.S. Children With ADHD, Autism, and Co-occurring ADHD-Autism: Evidence From the 2023 to 2024 National Survey of Children’s Health

**DOI:** 10.64898/2026.10.02.26364593

**Authors:** Chika Onwuameze, Godyson Orji, Martin Ayuk Bate, Vivian Ogechi Madu, Philip Kaya, Joy Inalegwu

**Author notes:** **Corresponding Author:** Chika Onwuameze, Ed.D., MPA Department of Public Administration Grambling State University, 403 Main Street, Grambling, Louisiana 71245 United States.

## Abstract

**Objective:** To compare healthcare access among U.S. children with attention-deficit/hyperactivity disorder (ADHD) only, autism only, and co-occurring ADHD–autism, and to identify social inequities associated with unmet care using recent nationally representative data.

**Methods:** We analyzed combined 2023 to 2024 National Survey of Children’s Health data for 13,576 children aged 3 to 17 years with current ADHD and/or autism. We examined six healthcare-access outcomes: forgone healthcare, unmet mental health care, no medical home, coverage gap/inadequate insurance, unmet specialist care, and unmet care coordination. Survey-weighted modified Poisson regression estimated adjusted prevalence ratios (aPRs) and 95% confidence intervals (CIs). We combined six multiply imputed poverty measures using Rubin’s rules. Sensitivity models additionally adjusted for functional difficulty.

**Results:** Compared with ADHD only, co-occurring ADHD–autism was associated with higher prevalence of forgone healthcare (aPR, 1.73; 95% CI, 1.38–2.16), no medical home (1.11; 1.02– 1.20), inadequate or discontinuous insurance (1.14; 1.03–1.28), unmet specialist care (1.49; 1.14–1.94), and unmet care coordination (1.15; 1.04–1.28). Autism only was associated with unmet mental health care (1.49; 1.13–1.97) and care coordination (1.13; 1.02–1.25). Food insufficiency, housing instability, and non-supportive neighborhood conditions were consistently associated with poorer access. After functional difficulty adjustment, associations remained for co-occurring ADHD–autism with forgone and specialist care and for autism only with unmet mental health care.

**Conclusions:** Healthcare access inequities differed by neurodevelopmental diagnostic group and social context. Pediatric systems should strengthen care coordination, specialty and mental health access, insurance continuity, family navigation, and responses to food and housing hardship.

## Introduction

Attention-deficit/hyperactivity disorder (ADHD) and autism spectrum disorder (autism) are among the most common neurodevelopmental conditions affecting U.S. children and adolescents. National estimates indicate that 10.5% of children aged 3-17 years had current ADHD in 2022, with most affected children having at least one co-occurring disorder.^1^ Recent analyses of the National Survey of Children’s Health (NSCH) have likewise documented autism prevalence near 3% among children and adolescents, with meaningful variation by sex, race and ethnicity, income, and insurance status.^2^ ADHD and autism also frequently co-occur. Children with both conditions have been shown to carry a greater burden of anxiety, depression, behavioral problems, and other developmental or mental health conditions than children with autism alone.^3^ This combination of needs can increase the complexity of care required across primary care, mental health, specialty, and community settings.

Greater clinical need does not necessarily translate into reliable access to services. A recent national payer analysis found that children with ADHD, autism, or both had markedly higher healthcare utilization and expenditures than children without these diagnoses.^4^ Autism also overlaps substantially with medical complexity, which can increase reliance on coordinated, multidisciplinary care.^5^ For autistic children, access to a medical home has been associated with fewer difficulties obtaining mental health and non-mental specialty services and lower unmet mental healthcare need.^6^ These findings underscore that continuity, coordination, specialty access, and insurance adequacy are central dimensions of healthcare quality for children with neurodevelopmental conditions, not simply measures of service use.

Access is also shaped by social and structural conditions. Among U.S. youth with common mental health problems, recent NSCH evidence identified persistent disparities in forgone preventive care and unmet mental healthcare after adjustment for income, parental education, and insurance.^7^ National geographic analyses have documented unequal access to autism resources across racially and ethnically diverse communities.^8^ Insurance design, cost sharing, health literacy, and family resources can further influence whether children obtain recommended specialty and supportive services. Recent work among Latinx families of autistic children illustrates how insurance type and health literacy may influence service access within an integrated healthcare system.^9^ Together, this literature suggests that poverty, insurance, food and housing insecurity, neighborhood conditions, and family educational resources may operate alongside diagnosis and functional need to shape access to care.

Important gaps remain. Much of the existing literature evaluates ADHD or autism separately, focuses on diagnosis or treatment rather than broad healthcare access, uses earlier survey periods, or relies on selected insured populations.^3,4,6^ Few recent nationally representative studies have directly compared children with ADHD only, autism only, and co-occurring ADHD-autism across multiple access domains while simultaneously accounting for socioeconomic, neighborhood, demographic, geographic, and functional factors. This distinction matters because co-occurring conditions may require more intensive coordination across primary, behavioral, specialty, and supportive services. Using combined 2023-2024 NSCH data, the objective of this study was to compare healthcare access among U.S. children aged 3-17 years with ADHD only, autism only, or co-occurring ADHD-autism and to identify social inequities associated with forgone healthcare, unmet mental health and specialist care, lack of a medical home, inadequate or discontinuous insurance, and unmet care coordination.

## Methods

### Study Design and Data Source

We conducted a cross-sectional analysis of the combined 2023-2024 National Survey of Children’s Health (NSCH). The NSCH is an annual, nationally representative household survey sponsored by the Health Resources and Services Administration’s Maternal and Child Health Bureau and administered by the U.S. Census Bureau. After an address-based household screener, one child aged 0-17 years is selected for a detailed age-specific topical questionnaire completed by a parent or caregiver knowledgeable about the child.^11^ The present study used the 2023-2024 combined topical Stata indicator file assembled by the Child and Adolescent Health Measurement Initiative Data Resource Center (CAHMI/DRC).^12^ The combined file contained 106,537 child records. Survey items, derived indicators, and coding followed CAHMI/DRC codebook and Census Bureau technical documentation.

### Study Population

The study population was restricted to children aged 3-17 years with a current parent-reported diagnosis of ADHD, autism, or both. Children younger than 3 years, those with neither current condition, and those without sufficient information to determine diagnostic status were excluded. The final analytic population included 13,576 children: 9,466 with ADHD only, 2,114 with autism only, and 1,996 with co-occurring ADHD and autism.

### Diagnostic Groups

Current ADHD and autism status were defined from the NSCH derived variables ADHD_2324 and autism_2324. Children identified as currently having a condition were classified as current; those reported as never having the condition or having been diagnosed previously but not currently having it were classified as not current. Three mutually exclusive groups were constructed: ADHD only, autism only, and ADHD + autism. ADHD only served as the reference group in regression analyses. This classification was intended to distinguish access patterns associated with each condition from those associated with co-occurrence, which has been shown to represent a clinically distinct population with greater comorbidity burden.^3^

### Healthcare Access Outcomes

Six binary healthcare-access outcomes were examined. Forgone healthcare identified children for whom needed healthcare was not received during the past 12 months; the comparison category combined children who received needed care and those for whom care was not needed. Unmet mental health care was defined among children who received mental health care or needed to see a mental health professional, with nonreceipt despite need coded as the adverse outcome. No medical home indicated that the child’s care did not meet NSCH medical-home criteria. Coverage gap or inadequate insurance distinguished children whose coverage was not both continuous and adequate from those with adequate, continuous coverage. Unmet specialist care was defined among children who received or needed specialist care, and unmet care coordination among children for whom care coordination was needed. These domains are consistent with healthcare-access constructs used in recent NSCH studies.^6,7^

The corresponding unweighted outcome-specific denominators were 13,525 for forgone healthcare, 7,145 for unmet mental health care, 13,566 for medical home, 13,494 for insurance adequacy/continuity, 4,335 for specialist care, and 9,998 for care coordination.

### Social and Demographic Measures

Social measures were selected a priori based on the NSCH framework and prior evidence linking social risks with pediatric chronic conditions and healthcare access.^7,10^ Household poverty was categorized as <100%, 100-199%, 200-399%, or >=400% of the federal poverty level (FPL). Highest adult education was categorized as less than high school, high school/GED, some college or technical school, or college degree or higher. Insurance type was classified as private only, public only, public and private, or uninsured. Food situation was categorized as enough nutritious food, enough food but not always nutritious, or sometimes/often not enough food. Housing instability was coded as experienced versus not experienced. Neighborhood safety was classified as definitely safe, somewhat safe, or not safe, and neighborhood support as supportive versus not supportive.

Demographic covariates included age group (3-5, 6-11, or 12-17 years), sex, race/ethnicity (Hispanic; non-Hispanic White; non-Hispanic Black; or other/multiracial non-Hispanic), 10-category HRSA region, and survey year. Reference categories were ADHD only, >=400% FPL, college degree or higher, private insurance, enough nutritious food, no housing instability, definitely safe neighborhood, supportive neighborhood, age 6-11 years, female sex, non-Hispanic White race/ethnicity, HRSA Region 1, and 2023. Insurance type was omitted from the insurance adequacy/continuity model because of conceptual overlap with the outcome.

### Functional Difficulty

Functional difficulty was categorized as none, one, or two or more functional difficulties. It was not included in the primary adjustment set because functional impairment may reflect clinical severity and may partly lie on the pathway between diagnostic group and healthcare need. Instead, functional difficulty was added in sensitivity analyses to assess whether diagnostic-group differences persisted after accounting for functional burden.

### Statistical Analysis

All analyses incorporated the complex NSCH sampling design. Survey strata were defined by crossing state with the within-state stratum variable; household identifier was specified as the cluster, and the CAHMI 2023-2024 combined child weight was applied. The full survey design was constructed before restricting to the ADHD/autism subpopulation so that variance estimation retained the parent sample structure. Multi-year weighting followed Census Bureau guidance for pooled-year estimation.^14^ Survey-weighted percentages and 95% confidence intervals were calculated for descriptive analyses.

Household poverty is multiply imputed in the NSCH using six FPL implicates.^13^ Each poverty estimate and each multivariable model was therefore estimated separately across all six implicates. Regression coefficients and design-based variances were combined using Rubin’s rules so that uncertainty from imputation was incorporated into standard errors and confidence intervals.^13^ Other missing covariate data were handled by complete-case analysis within each outcome-specific model; no additional imputation was performed.

Because several outcomes were common, associations were estimated using survey-weighted modified Poisson regression with a log link and design-based robust variance, yielding adjusted prevalence ratios (aPRs) and 95% confidence intervals rather than odds ratios.^15^ Analyses were conducted in R using methods for stratified, clustered, and weighted survey data.^16^ Primary models included diagnostic group, social measures, demographic characteristics, HRSA region, and survey year; sensitivity models additionally included functional difficulty.

Interpretation emphasized aPRs and 95% confidence intervals. The prespecified objective was to determine whether healthcare-access inequities differed among children with ADHD only, autism only, and co-occurring ADHD-autism after accounting for social and demographic factors, and whether those differences persisted after additional adjustment for functional difficulty.

### Ethics

The analysis used publicly available, de-identified NSCH data and involved no direct contact with participants. [Insert the institutional review board determination or exemption statement required by the submitting institution and journal.]

## Results

### Study Population

The analytic sample included 13,576 children aged 3-17 years with current ADHD and/or autism in the combined 2023-2024 National Survey of Children’s Health. Of these, 9,466 had ADHD only, 2,114 had autism only, and 1,996 had co-occurring ADHD and autism. After survey weighting, these groups represented 67.5%, 17.2%, and 15.4% of the study population, respectively. Children with autism only and those with co-occurring ADHD-autism were more likely than children with ADHD only to be male and to experience socioeconomic disadvantage. Housing instability affected 22.5% of the ADHD-only group, 28.8% of the autism-only group, and 30.1% of the co-occurring group. Functional burden differed markedly: two or more functional difficulties were present in 26.2%, 37.5%, and 52.6%, respectively (Table 1).

**Table 1.** Selected characteristics of children with ADHD and/or autism, NSCH 2023-2024.

| **Characteristic** | **ADHD only** | **Autism only** | **ADHD +**<br>**autism** |
| --- | --- | --- | --- |
| Unweighted n | 9,466 | 2,114 | 1,996 |
| Weighted share of analytic population, % | 67.5 | 17.2 | 15.4 |
| Male, % | 62.5 | 72.0 | 73.9 |
| Household income <100% FPL, % | 18.4 | 21.4 | 23.1 |
| Housing instability, % | 22.5 | 28.8 | 30.1 |
| Two or more functional difficulties, % | 26.2 | 37.5 | 52.6 |

FPL, federal poverty level. Percentages are survey weighted.

### Healthcare Access Outcomes

Weighted healthcare-access outcomes differed across diagnostic groups (Table 2). Forgone healthcare was reported for 7.7% of children with ADHD only, 10.5% with autism only, and 15.5% with co-occurring ADHD-autism. Among children who received or needed mental health care, unmet need affected 14.5%, 24.7%, and 18.0%, respectively. Care did not meet medical-home criteria for 57.4%, 68.5%, and 67.1%. Coverage gaps or inadequate insurance affected 38.1%, 36.9%, and 43.2%. Among children who received or needed specialist care, unmet specialist care was highest in the co-occurring group at 27.1%, compared with 14.5% for ADHD only and 15.6% for autism only. Unmet care coordination affected 44.0%, 55.3%, and 53.4%, respectively.

**Table 2.** Weighted prevalence of primary healthcare-access outcomes.

| **Outcome** | **ADHD only,**<br>**%** | **Autism only,**<br>**%** | **ADHD +**<br>**autism, %** |
| --- | --- | --- | --- |
| Forgone healthcare | 7.7 | 10.5 | 15.5 |
| Unmet mental health care* | 14.5 | 24.7 | 18.0 |
| No medical home | 57.4 | 68.5 | 67.1 |
| Coverage gap/inadequate insurance | 38.1 | 36.9 | 43.2 |
| Unmet specialist care† | 14.5 | 15.6 | 27.1 |
| Unmet care coordination‡ | 44.0 | 55.3 | 53.4 |

\*Among children who received or needed mental health care.
†Among children who received or needed specialist care.
‡Among children who needed care coordination.

### Adjusted Associations

In fully adjusted survey-weighted modified Poisson models, diagnostic-group differences varied by access domain (Table 3; Figure 1). Compared with ADHD only, co-occurring ADHD-autism was associated with greater prevalence of forgone healthcare (aPR, 1.73; 95% CI, 1.38-2.16), whereas autism only was not (aPR, 1.21; 95% CI, 0.91-1.61). Forgone healthcare was also more prevalent among uninsured children (aPR, 1.93; 95% CI, 1.26-2.94), children in households that sometimes or often lacked enough food (aPR, 1.99; 95% CI, 1.39-2.84), those experiencing housing instability (aPR, 1.45; 95% CI, 1.16-1.82), those living in neighborhoods rated not safe (aPR, 1.55; 95% CI, 1.11-2.15), and those in non-supportive neighborhoods (aPR, 2.00; 95% CI, 1.58-2.53). Forgone healthcare was less prevalent in 2024 than in 2023 (aPR, 0.78; 95% CI, 0.65-0.93).

**Figure 1.**
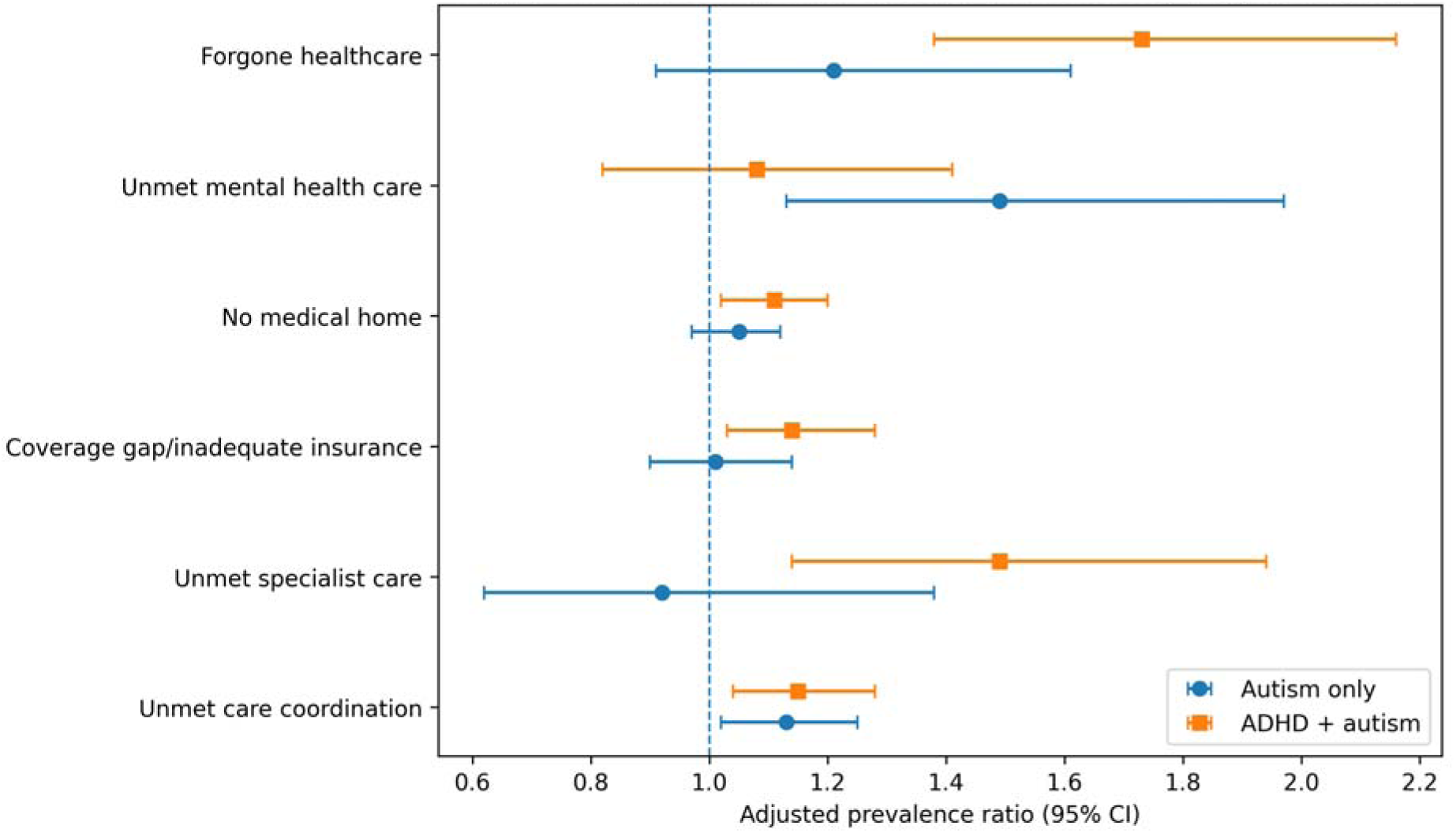
Adjusted healthcare-access differences by diagnostic group. Forest plot of adjusted prevalence ratios (aPRs) and 95% confidence intervals for autism only and co-occurring ADHD-autism, with ADHD only as the reference group. The dashed vertical line denotes an aPR of 1.00.

**Table 3.** Primary adjusted diagnostic-group associations with healthcare-access outcomes.

| **Outcome** | **Autism only vs ADHD**<br>**only, aPR (95% CI)** | **ADHD + autism vs**<br>**ADHD only, aPR (95%**<br>**CI)** |
| --- | --- | --- |
| Forgone healthcare | 1.21 (0.91-1.61) | 1.73 (1.38-2.16) |
| Unmet mental health care | 1.49 (1.13-1.97) | 1.08 (0.82-1.41) |
| No medical home | 1.05 (0.97-1.12) | 1.11 (1.02-1.20) |
| Coverage gap/inadequate insurance | 1.01 (0.90-1.14) | 1.14 (1.03-1.28) |
| Unmet specialist care | 0.92 (0.62-1.38) | 1.49 (1.14-1.94) |
| Unmet care coordination | 1.13 (1.02-1.25) | 1.15 (1.04-1.28) |

Note: Models additionally adjusted for poverty, adult education, insurance type, food situation, housing instability, neighborhood safety, neighborhood support, age, sex, race/ethnicity, HRSA region, and survey year. Insurance type was omitted from the insurance adequacy/continuity model. Six FPL implicates were combined using Rubin's rules.

For unmet mental health care, autism only was associated with higher prevalence than ADHD only (aPR, 1.49; 95% CI, 1.13-1.97), whereas the estimate for co-occurring ADHD-autism was not elevated (aPR, 1.08; 95% CI, 0.82-1.41). Uninsurance (aPR, 1.69; 95% CI, 1.12-2.55), sometimes or often insufficient food (aPR, 2.04; 95% CI, 1.37-3.04), housing instability (aPR, 1.39; 95% CI, 1.08-1.77), and a non-supportive neighborhood (aPR, 1.41; 95% CI, 1.11-1.79) were also associated with greater unmet mental health need.

Co-occurring ADHD-autism was associated with higher prevalence of not meeting medical-home criteria (aPR, 1.11; 95% CI, 1.02-1.20) and coverage gaps or inadequate insurance (aPR, 1.14; 95% CI, 1.03-1.28). In the insurance adequacy model, sometimes or often insufficient food (aPR, 1.38; 95% CI, 1.17-1.62), somewhat unsafe neighborhoods (aPR, 1.15; 95% CI, 1.05-1.27), and non-supportive neighborhoods (aPR, 1.24; 95% CI, 1.13-1.36) were associated with higher prevalence.

The largest diagnostic-group difference in specialist access occurred among children with co-occurring ADHD-autism, who had 49% higher adjusted prevalence of unmet specialist care than children with ADHD only (aPR, 1.49; 95% CI, 1.14-1.94). Compared with >=400% of the federal poverty level (FPL), aPRs were 1.92 (95% CI, 1.16-3.20) for 100%-199% FPL and 1.97 (95% CI, 1.13-3.43) for <100% FPL. Lower adult education showed a gradient, reaching an aPR of 2.31 (95% CI, 1.38-3.88) for less than high school education. Housing instability was also associated with unmet specialist care (aPR, 1.44; 95% CI, 1.06-1.97).

Both autism only (aPR, 1.13; 95% CI, 1.02-1.25) and co-occurring ADHD-autism (aPR, 1.15; 95% CI, 1.04-1.28) were associated with unmet care coordination. Sometimes or often insufficient food (aPR, 1.41; 95% CI, 1.21-1.64), a neighborhood rated not safe (aPR, 1.23; 95% CI, 1.08-1.41), and lack of neighborhood support (aPR, 1.38; 95% CI, 1.26-1.51) were also associated with unmet coordination.

### Sensitivity Analyses

Additional adjustment for functional difficulty changed several diagnostic-group estimates (Table 4; Figure 2). The association between co-occurring ADHD-autism and forgone healthcare attenuated but remained elevated (aPR, 1.42; 95% CI, 1.14-1.77), as did the association with unmet specialist care (aPR, 1.48; 95% CI, 1.13-1.93). The autism-only association with unmet mental health care was essentially unchanged (aPR, 1.48; 95% CI, 1.12-1.96). In contrast, the co-occurring-group associations with no medical home (aPR, 1.05; 95% CI, 0.97-1.14), coverage gap/inadequate insurance (aPR, 1.09; 95% CI, 0.98-1.23), and unmet care coordination (aPR, 1.05; 95% CI, 0.95-1.17) attenuated after functional-difficulty adjustment. The autism-only estimate for unmet care coordination similarly attenuated to 1.09 (95% CI, 0.98-1.20). Overall, the analyses identified diagnostic-group and social inequities across six healthcare-access domains and showed which differences persisted after accounting for functional burden.

**Figure 2.**
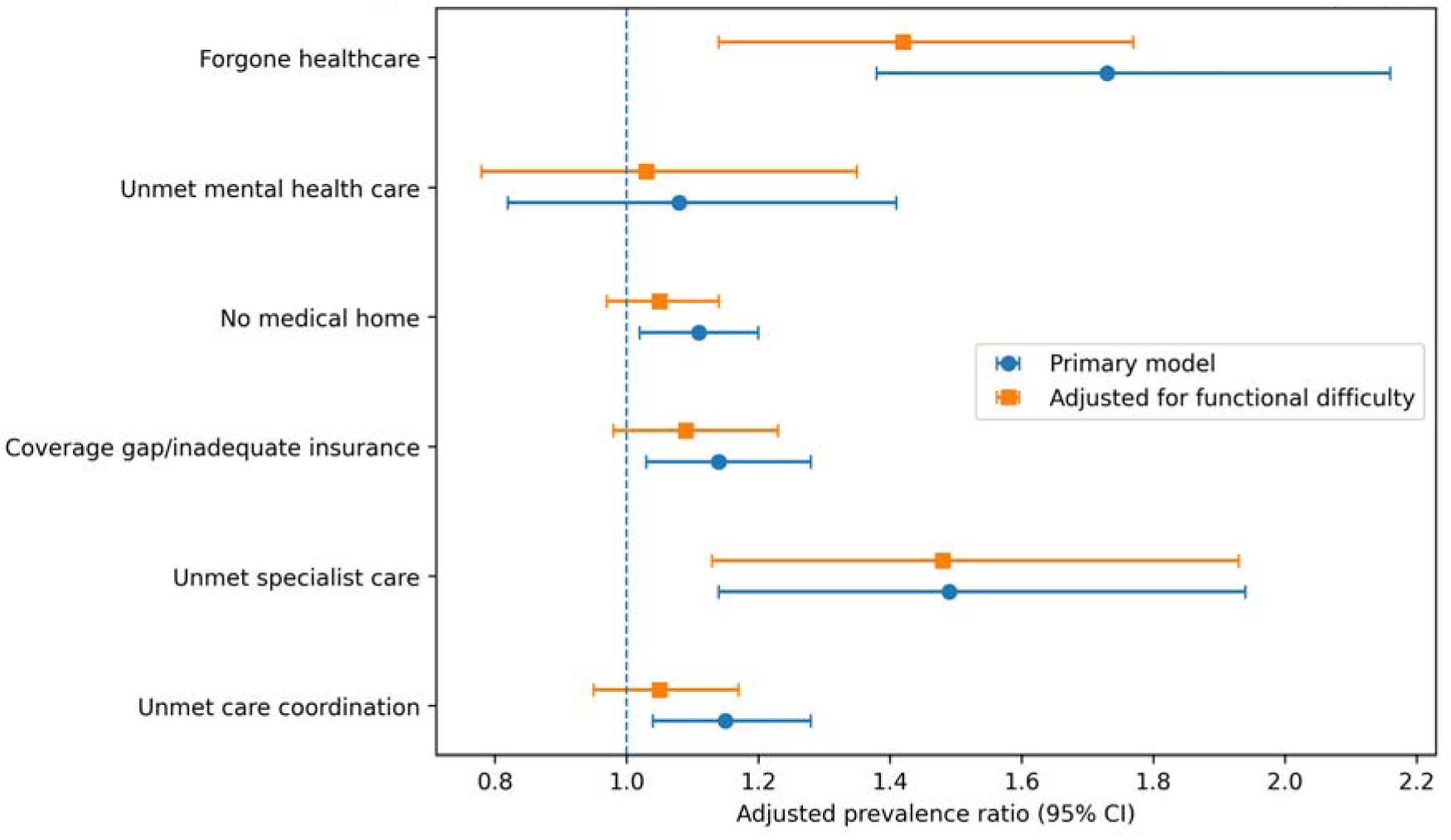
Sensitivity of ADHD + autism associations to additional adjustment for functional difficulty. Forest plot comparing primary and functional-difficulty-adjusted aPRs and 95% confidence intervals across the six healthcare-access outcomes. The dashed vertical line denotes an aPR

**Table 4.** Sensitivity analysis after additional adjustment for functional difficulty.

| **Outcome** | **Group** | **Primary aPR**<br>**(95% CI)** | **Functional-difficulty-**<br>**adjusted aPR (95%**<br>**CI)** |
| --- | --- | --- | --- |
| Forgone healthcare | Autism only | 1.21 (0.91-1.61) | 1.12 (0.83-1.51) |
|  | ADHD + autism | 1.73 (1.38-2.16) | 1.42 (1.14-1.77) |
| Unmet mental health care | Autism only | 1.49 (1.13-1.97) | 1.48 (1.12-1.96) |
|  | ADHD + autism | 1.08 (0.82-1.41) | 1.03 (0.78-1.35) |
| No medical home | Autism only | 1.05 (0.97-1.12) | 1.03 (0.96-1.10) |
|  | ADHD + autism | 1.11 (1.02-1.20) | 1.05 (0.97-1.14) |
| Coverage gap/inadequate insurance | Autism only | 1.01 (0.90-1.14) | 1.00 (0.88-1.13) |
|  | ADHD + autism | 1.14 (1.03-1.28) | 1.09 (0.98-1.23) |
| Unmet specialist care | Autism only | 0.92 (0.62-1.38) | 0.92 (0.61-1.38) |
|  | ADHD + autism | 1.49 (1.14-1.94) | 1.48 (1.13-1.93) |
| Unmet care coordination | Autism only | 1.13 (1.02-1.25) | 1.09 (0.98-1.20) |
|  | ADHD + autism | 1.15 (1.04-1.28) | 1.05 (0.95-1.17) |

## Discussion

Healthcare access differed across children with ADHD only, autism only, and co-occurring ADHD-autism, and the pattern varied by type of care. Children with co-occurring ADHD-autism had higher adjusted prevalence of forgone healthcare, unmet specialist care, lack of a medical home, inadequate or discontinuous insurance, and unmet care coordination than children with ADHD only, whereas children with autism only had higher unmet mental health care and care-coordination needs. These findings extend evidence that children with autism, ADHD, or both have high healthcare utilization and expenditures and that autism often overlaps with medical complexity.^4,5^ Greater need did not ensure that needed services were obtained.

Diagnostic-group differences were particularly pronounced for forgone healthcare and specialist care. After adjustment for socioeconomic, demographic, geographic, and neighborhood factors, children with co-occurring ADHD-autism had 73% higher prevalence of forgone healthcare and 49% higher prevalence of unmet specialist care than children with ADHD only (Table 3; Figure 1). This pattern is consistent with greater comorbidity among children with co-occurring neurodevelopmental conditions.^3,4^ Recent NSCH research has also identified barriers to autism-related behavioral therapy, while medical-home care in autism has been associated with less difficulty obtaining mental and nonmental specialty services.^6,17^ Together, these findings suggest a mismatch between required services and the capacity of families and health systems to coordinate them.

Social conditions were independently associated with access across several outcomes. Food insufficiency was linked to forgone healthcare, unmet mental health care, inadequate insurance, lack of a medical home, and unmet care coordination; housing instability was associated with forgone care, mental health care, and specialist access; and lack of neighborhood support was consistently associated with poorer access. These patterns align with evidence that unstable housing clusters with food-related and healthcare hardships and that lower-opportunity neighborhoods are associated with poorer healthcare experiences.^18,21^ National NSCH data have also linked limited community support with unmet healthcare needs.^19^ A systematic review of ADHD found a socioeconomic gradient in service use and health-related quality of life, supporting the role of social resources in translating clinical need into care.^20^

The findings for mental health and specialist care warrant particular attention. Autism only was associated with approximately 49% higher adjusted prevalence of unmet mental health care, and this estimate was essentially unchanged after adjustment for functional difficulty. This is consistent with recent national evidence of substantial unmet mental healthcare among U.S. youth with behavioral and emotional conditions.^7^ For specialist care, household income below 200% of the federal poverty level and lower adult education were associated with markedly higher unmet need, while the co-occurring ADHD-autism association persisted. Recent autism studies likewise show that insurance, caregiver education, health literacy, geography, and resource availability influence access to recommended services.^8,9,17^ These results support interventions that address navigation and affordability as well as specialist supply.

The sensitivity analyses clarify the role of functional burden (Table 4; Figure 2). Adjustment for functional difficulty attenuated the co-occurring ADHD-autism associations with medical-home status, insurance adequacy, and care coordination, and the autism-only association with care coordination. In contrast, associations of co-occurring ADHD-autism with forgone healthcare and unmet specialist care, and of autism only with unmet mental health care, remained elevated. Functional difficulty therefore explained part, but not all, of the diagnostic-group differences. Recent national work shows extensive overlap between autism and medical complexity, including functional limitations and high healthcare expenditures.^5^ The persistence of several associations after severity adjustment suggests that access barriers cannot be attributed solely to greater clinical need.

These findings have practical implications for pediatric care. Practices caring for children with ADHD or autism should identify co-occurrence explicitly and assess access needs alongside symptoms and functional impairment. For children with co-occurring conditions, active referral tracking, shared care plans, closed-loop communication with mental health and specialty clinicians, and family navigation should replace reliance on families to coordinate multiple systems independently. Medical-home models are especially relevant because coordinated care has been associated with fewer difficulties obtaining mental and specialty services among autistic children.^6^ Practices should also screen for food and housing hardship and connect families with benefits, community organizations, and social-service partners.

Policy responses should be specific. States and health plans can strengthen continuous Medicaid and CHIP enrollment, simplify renewal processes, reimburse care coordination and family-navigation services, and expand telehealth and specialty networks where autism resources are limited. Autism services are unevenly distributed, with resource gaps affecting some racial and ethnic communities.^8^ Recent work also supports telehealth expansion, insurance outreach, and family navigation.^17^ The lower prevalence of inadequate or discontinuous insurance observed in some lower-income groups may reflect public coverage eligibility and should not be interpreted as poverty being protective; rather, it highlights the potential buffering role of public insurance while other access barriers persist.

This study has several strengths. It used recent 2023-2024 nationally representative data; distinguished ADHD only, autism only, and co-occurring ADHD-autism; evaluated six clinically relevant access outcomes; preserved the complex survey structure for subpopulation analysis; incorporated all six multiply imputed poverty implicates; and estimated prevalence ratios appropriate for common outcomes. The functional-difficulty sensitivity analyses further helped distinguish differences related to clinical need from broader access inequities.

Several limitations warrant consideration. The cross-sectional design precludes causal inference and temporal ordering. Diagnoses, healthcare need, and access were parent reported and may be affected by recall or recognition of need. The forgone-care comparison category combined children who received needed care with those who did not need care, complicating interpretation of some covariate associations. Residual confounding is possible, and covariates other than poverty were handled using complete-case analysis. HRSA regions may also mask substantial state and local variation in workforce availability, rurality, benefit design, and service supply.

Using nationally representative 2023-2024 NSCH data, this study found meaningful healthcare-access differences among children with ADHD only, autism only, and co-occurring ADHD-autism. Children with co-occurring conditions experienced particularly high prevalence of forgone healthcare and unmet specialist care, while children with autism alone had greater unmet mental healthcare need. Several disparities persisted after accounting for functional difficulty, indicating that clinical complexity alone does not explain unequal access. Pediatric systems should prioritize closed-loop specialty referrals, mental health integration, family navigation, continuous insurance coverage, and systematic screening and referral for food, housing, and neighborhood-related needs.^5,17,18^ These actions directly address the study objective of identifying modifiable healthcare-access inequities across neurodevelopmental diagnostic groups.

## Data Availability

The data used in this study were obtained from the 2023 to 2024 National Survey of Childrens Health through the Child and Adolescent Health Measurement Initiative Data Resource Center. The original NSCH data are available from the official data source subject to the applicable access terms. Analytic code, variable definitions, derived study variables, and supporting materials will be archived in Zenodo. The original NSCH microdata are not redistributed by the authors.

https://www.childhealthdata.org/help/dataset/2023-2024-combined-nsch-dataset---codebook-instruction

https://www.census.gov/programs-surveys/nsch/data/datasets.html

